# Efficacy and safety of non-invasive ventilation in pediatric care in low-income and middle-income countries: a systematic review

**DOI:** 10.1101/2021.07.27.21261207

**Authors:** Kristen L Sessions, Andrew Gerald Smith, Peter J Holmberg, Tisungane Mvalo, Mohammod Jobayer Chisti, Ryan W. Carroll, Eric D McCollum

**Affiliations:** Department of Pediatrics, Ann & Robert H. Lurie Children’s Hospital of Chicago, Chicago, IL, USA; Division of Pediatric Critical Care Medicine, University of Utah, Salt Lake City, Utah, USA; Division of Pediatric Hospital Medicine, Department of Pediatric and Adolescent Medicine, Mayo Clinic Children’s Center, Rochester, MN, USA; Department of Pediatrics, University of North Carolina Project Malawi, Lilongwe, Malawi, University of North Carolina, Chapel Hill, NC, USA; International Centre for Diarrhoeal Disease Research, Dhaka, Bangladesh; Division of Pediatric Critical Care Medicine, MassGeneral Hospital for Children, Harvard School of Medicine, Boston, MA, USA; Johns Hopkins Global Program in Respiratory Sciences, Eudowood Division of Pediatric Respiratory Sciences, Department of Pediatrics, Johns Hopkins School of Medicine, Baltimore, USA; Department of International Health, Johns Hopkins Bloomberg School of Public Health, Baltimore, USA

## Abstract

**Background:** Lower respiratory tract infections (LRTIs) are a leading cause of under-5 mortality in low-income and middle-income countries (LMICs) and interventions to reduce mortality are needed. Non-invasive ventilation has been shown to reduce mortality for neonates; however, data for children >1 month of age in LMICs are lacking. The objective of this study was to systematically review the available literature to determine if non-invasive ventilation as the primary modality of respiratory support is efficacious and safe for the management of respiratory distress in non-neonatal pediatric patients in LMICs.

**Methods:** We systematically reviewed all studies assessing the endpoints of efficacy, effectiveness, and safety of non-invasive ventilation for pediatric LRTIs in LMICs. A comprehensive search of Medline, Embase, LILACS, Web of Science, and Scopus was performed on April 7, 2020. Included studies assessed the safety, efficacy or effectiveness of non-invasive ventilation (NIV) in the hospital setting for pediatric patients with respiratory distress from 1 month - 15 years of age in LMICs. All study types, including case reports and case series were included. Studies focusing exclusively on neonates (<28 days old) were excluded. Mortality and rates of adverse events were extracted using Covidence by two independent reviewers. Risk of bias was assessed using GRADE criteria for randomized control trials and a standardized risk of bias assessment tool for observational studies. The study protocol was registered on PROSPERO (CRD42018084278).

**Findings:** A total of 2174 papers were screened and 20 met criteria for inclusion. There were 5 randomized control trials (RCTs), including 3 large, well-designed RCTs. The first RCT, the ‘Bangladesh trial,’ found that children who received bubble continuous positive airway pressure (bCPAP) compared to low-flow oxygen had a significantly lower risk of failure (6% in CPAP and 24% in low-flow oxygen, p=0.0026) and mortality (4% in CPAP and 15% in low-flow oxygen, p=0.022). A second RCT, the ‘Ghana trial,’ found no decrease in all-cause mortality between the CPAP and control arms (3% and 4% respectively, p=0.11); however, an adjusted secondary analysis demonstrated decreased mortality for children under 1 year of age (3% in CPAP and 7% in control group, p=0.01). The third RCT, the ‘Malawi trial,’ compared bCPAP to low flow oxygen and found higher mortality in the bCPAP arm (17% and 11% respectively, p=0.036). Among the non-RCT studies, mortality rates ranged from 0-55%.

**Interpretation:** The evidence of efficacy, effectiveness, and safety is mixed regarding the use of NIV in children with respiratory failure in LMICs. Our review of the literature suggests that CPAP for non-neonatal pediatric patients should be considered only in well-controlled, high acuity units with high provider-to-patient ratios and direct physician supervision. Until further data are available, CPAP use in LMICs should be limited to children less than 1 year of age. Further research is needed to determine best practices for CPAP prior to wide-spread implementation.

**Funding:** There was no funding source for this study.

## Introduction

Significant progress has been made in reducing the global burden of mortality for children during the last twenty years, with overall rates decreasing by more than half since 2000. Despite these improvements, nearly 6.5 million children worldwide died in 2017, including 5.4 million under the age of 5 years.^1^ Reflecting historical trends, lower respiratory tract infections (LRTIs) continue to play a disproportionate role in child mortality, accounting for more deaths in children age 1-59 months than any other illness.^1^ Various efforts, including the formation of WHO treatment guidelines, a focus on child health in the Millennium Development Goals, and the subsequent advancement of the Sustainable Development Goals have led to dramatic reductions in child mortality secondary to LRTIs.^1^ However, large disparities persist, and mortality from respiratory causes is especially striking in low- and middle-income countries (LMICs).^2^

Current management techniques for LRTIs and respiratory distress generally include medical therapies such as intravenous fluids and antimicrobials, in addition to respiratory support. In many LMICs, the highest level of respiratory support available is often low-flow oxygen (typically 1-2 liters per minute, LPM) delivered by a simple nasal cannula or face mask. Some larger hospitals may have capacity for more intensive management, including non-invasive ventilation (NIV) and intubation with mechanical ventilation (IMV), but the necessary equipment, medications and high-level of care required makes this rare.

NIV provides positive airway pressure to a spontaneously breathing individual, helping maximize lung function and reduce ventilation-perfusion mismatch, thereby improving gas exchange and alleviate work of breathing.^3^ In high-income countries, NIV has become a standard of care for pediatric patients with respiratory distress or failure and can reduce rates of morbidity – reduce need for intubation and mechanical ventilation – and decrease mortality. A common form of NIV used worldwide is bubble continuous positive airway pressure, or ‘bubble CPAP’ (bCPAP). In LMICs, the use of bCPAP has been shown to be particularly beneficial in managing neonatal respiratory distress (<28 days old). A systematic review of bCPAP for neonates in LMICs demonstrated a reduction in the need for mechanical ventilation by 30-50% with no change in mortality.^4^ Safety concerns for NIV include possible excessive oxygen delivery, skin and/or nasal septal damage, and, rarely, a risk of pneumothorax.

While bCPAP for neonates in LMICs has been shown to be beneficial, the efficacy, effectiveness, and safety of NIV for non-neonatal pediatric patients in LMICs has been a recent focus. A systematic review of the literature through 2018 concluded that bCPAP was thus far safe and effective in LMICs.^5^ However, new research has been published in the subsequent years, raising new questions regarding NIV for non-neonates. The objective of this study was to systematically review the available literature to determine if NIV as the primary modality of respiratory support is efficacious, effective, and safe for the management of respiratory distress in pediatric patients 1 month – 15 years of age in LMICs. Secondary objectives were to define morbidity, treatment failure, and adverse events associated with the use of NIV in this target population.

## Methods

The development and reporting of this systematic review is based on the Preferred Reporting Items for Systematic Reviews (PRISMA) statement.^6^ The protocol was registered on PROSPERO (CRD42018084278).

### Data Sources and Search Strategies

A comprehensive search of Medline, Embase, LILACS, Web of Science, and Scopus was performed on April 7, 2020 based on the population, intervention, comparison, and outcome format (Table 1). There were no restrictions by language, publication date, or publication type. There were also no restrictions by age of participants as to not inadvertently exclude eligible studies. Countries of interest were defined by the World Bank classification of LMIC. The search strategy was designed and conducted by a medical reference librarian with input from the investigators (Appendix 1). The references of included studies were also searched.

**Table 1:** Search Criteria for Studies

| PICO Term | Description |
| --- | --- |
| Population | Patients 1 month – 15 years of age with respiratory distress including, but not limited to, pneumonia or bronchiolitis in low- and middle-income countries |
| Intervention | Non-invasive ventilation including bubble continuous positive airway pressure (bCPAP), positive end-expiratory pressure, and nasal continuous airway pressure (nCPAP) used in the acute hospital setting for treatment of respiratory distress |
| Comparison | High or low-flow oxygen therapy through nasal cannula, mechanical ventilation, or no respiratory support |
| Outcome | Mortality, morbidity, need for mechanical ventilation, treatment failure, adverse events |

### Inclusion and Exclusion Criteria

All studies published in peer-reviewed journals with a primary focus on the efficacy, effectiveness, or safety of NIV in the population of interest, including case reports and case series, were included. NIV was defined as bCPAP, CPAP, or nCPAP. Editorials, letters, narratives, systematic reviews, and errata were excluded. Included studies assessed the efficacy, effectiveness, or safety of NIV in the hospital setting for pediatric patients with respiratory distress from 1 month - 15 years of age in LMICs. Studies focusing exclusively on neonates (<28 days old) were excluded though studies including both neonates and children >1 month of age were included.

### Data Collection and Extraction

Search keywords are listed in Appendix 1. Sources were collected using EndNote and all data was managed using Covidence, an online platform for data extraction and quality assessments for systematic reviews. Each study was screened by title and abstract by two independent reviewers to assess for possible inclusion. Eligible studies underwent a full text review for final inclusion. Disagreements at the title and abstract review stage were resolved by a third blinded author while disagreements at the manuscript review stage were resolved by consensus discussion. A standard data extraction tool was created in Covidence. Extracted data included authors, funding, setting, study design and population, interventions, and outcomes including mortality and adverse events.

### Risk of Bias Assessment

Included papers were evaluated for risk of bias. Comparative studies, including all randomized control trails, were evaluated using the Cochrane GRADE Criteria, which evaluates sequence generation, allocation concealment, blinding, incomplete outcome data, and selective outcome reporting.^7^ Studies with no comparator group including retrospective and prospective observational studies, case series, and case studies were evaluated using the criteria proposed by Murad et.al. which evaluates the domains of selection, ascertainment, causality, and reporting.^8^ Data extraction and risk of bias assessments were performed by two independent reviewers and discrepancies were adjudicated by consensus discussion.

## Results

A total of 2174 studies were screened and 20 met criteria for inclusion (Figure 1). These included five randomized control trials (RCTs)^9-13^, one cluster RCT^14^, one non-randomized comparative study^15^, eleven observational studies^16-26^ (prospective and retrospective observational studies and case series) and two case reports^27,28^ (Table 2). A majority of the studies evaluated the use of NIV in the form of bubble CPAP (bCPAP) or conventional nasal CPAP (nCPAP). Other forms of NIV included conventional oropharyngeal CPAP and pressure controlled portable ventilation. Most studies were small with twelve studies enrolling less than 100 patients. Half the studies (10/20) included, but were not limited to, neonates (<1 month of age) in their analyses.

**Table 2:**
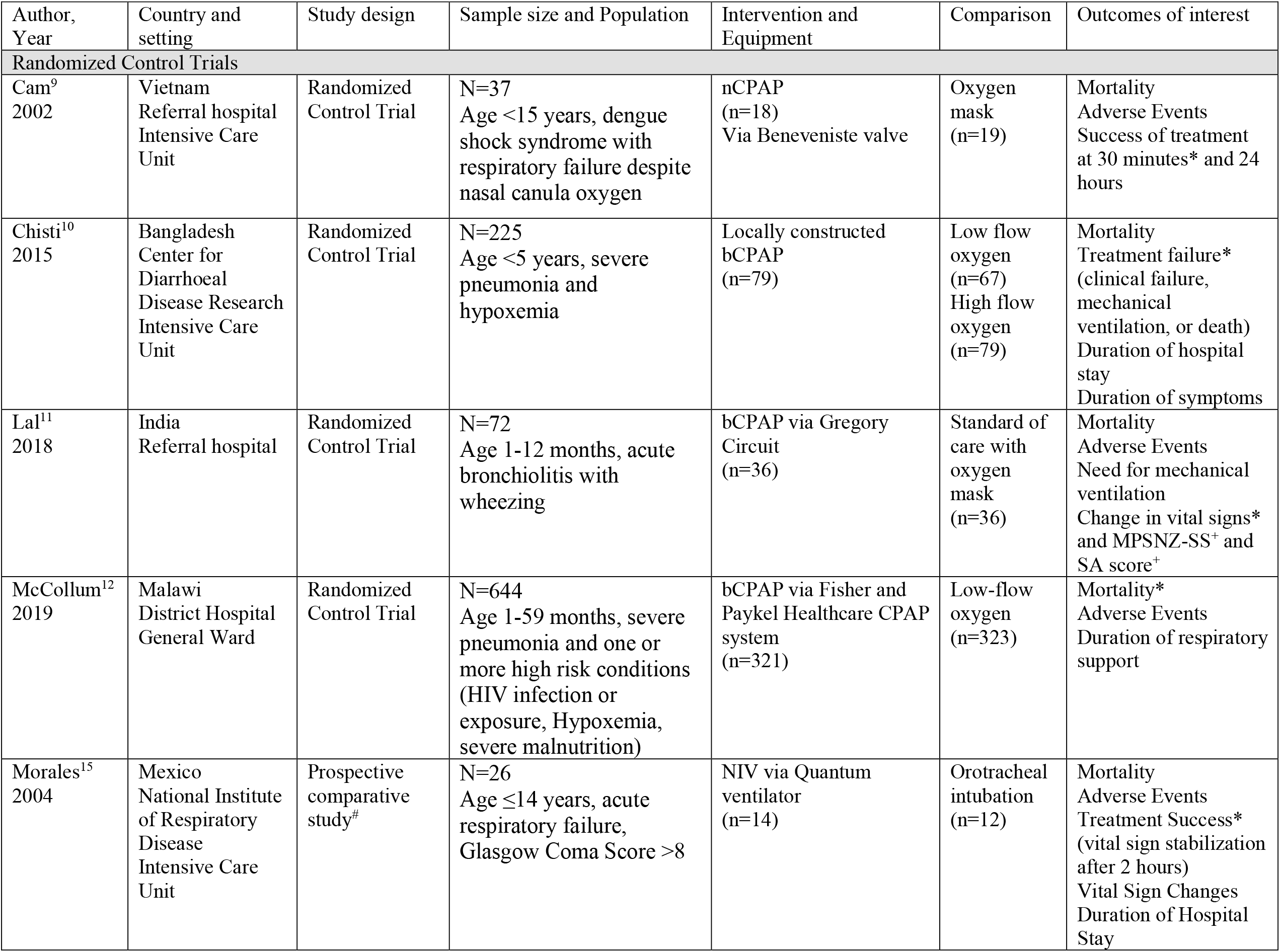
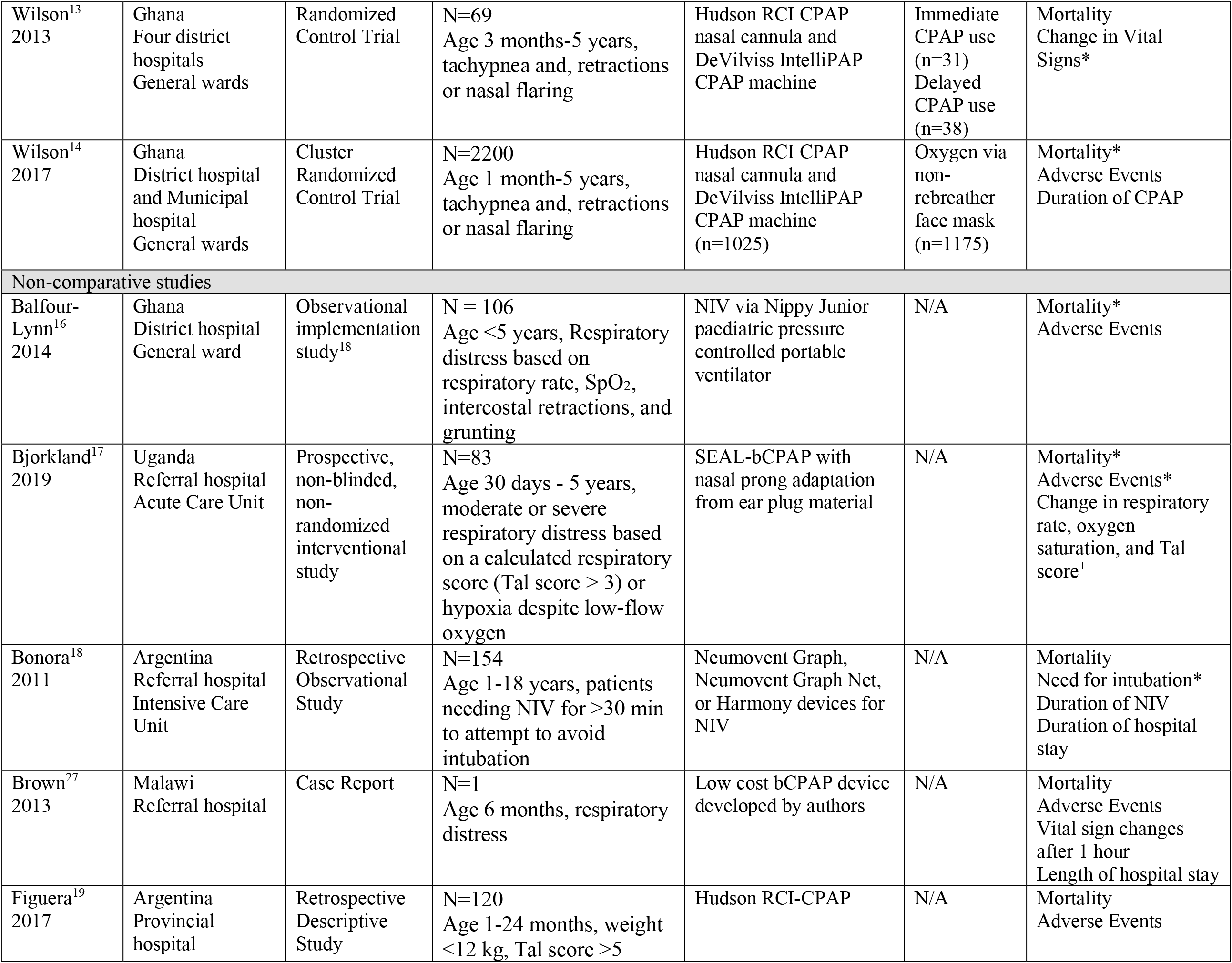
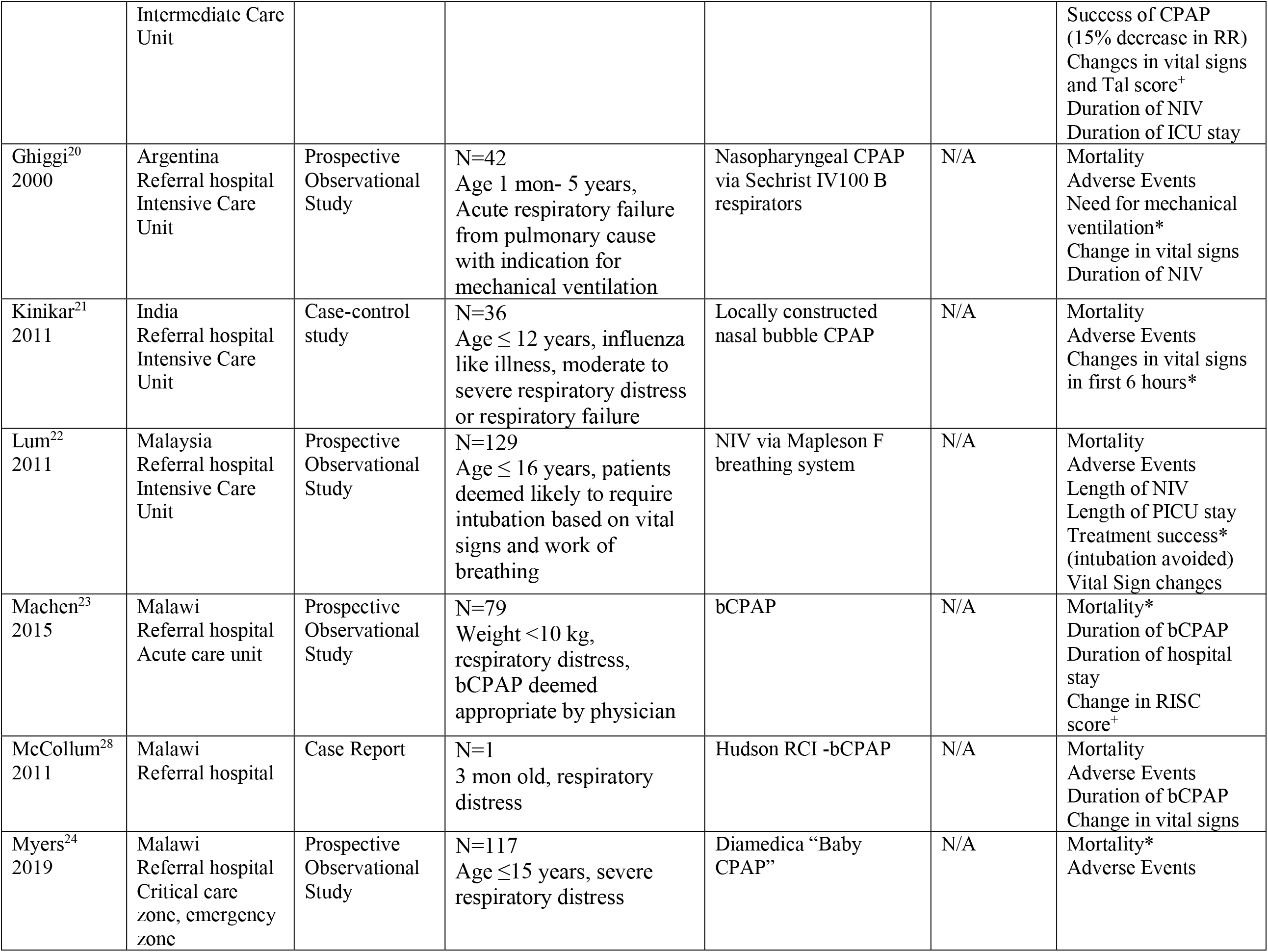
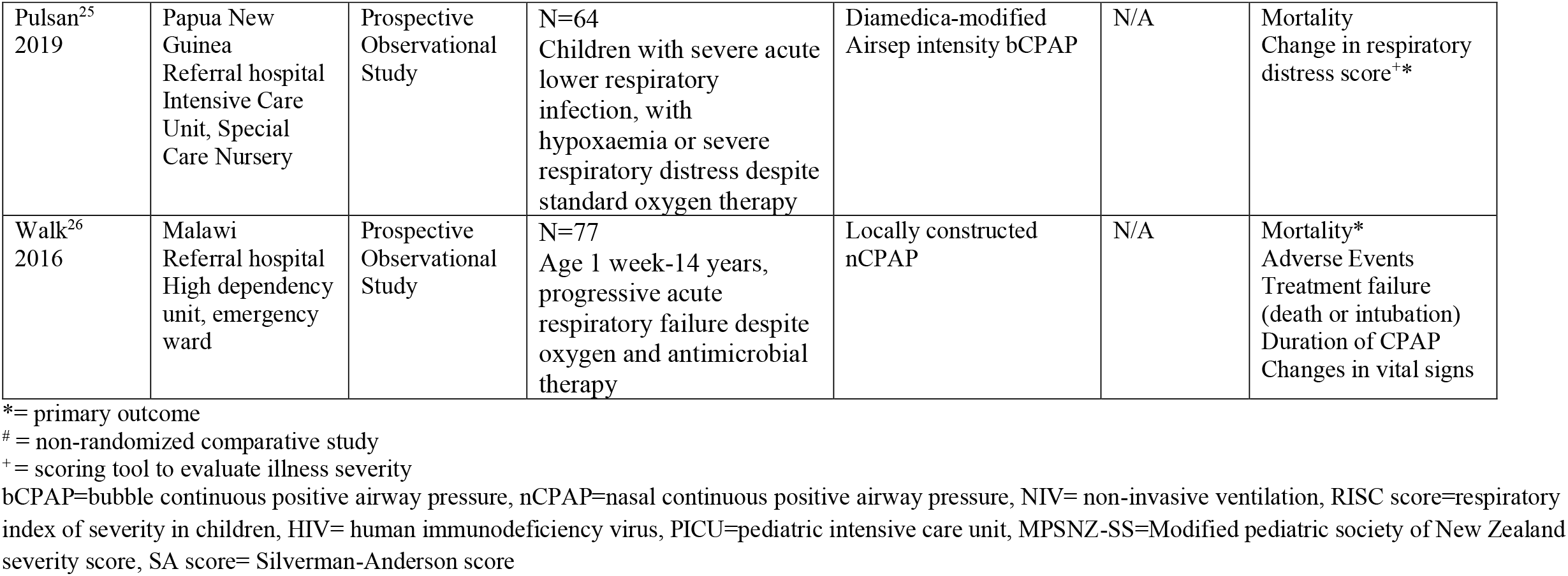
Characteristics of included studies

**Table 3a:**
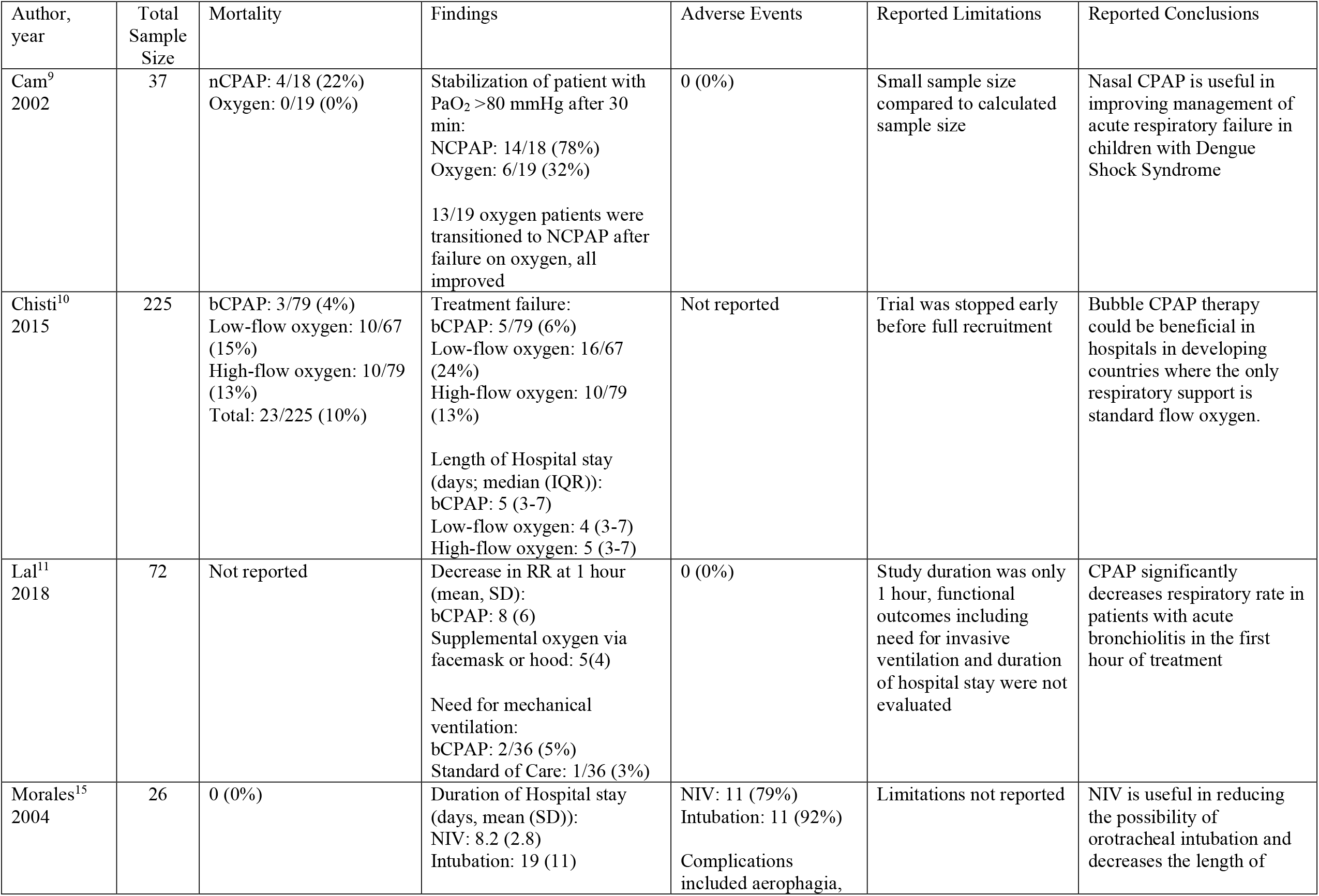
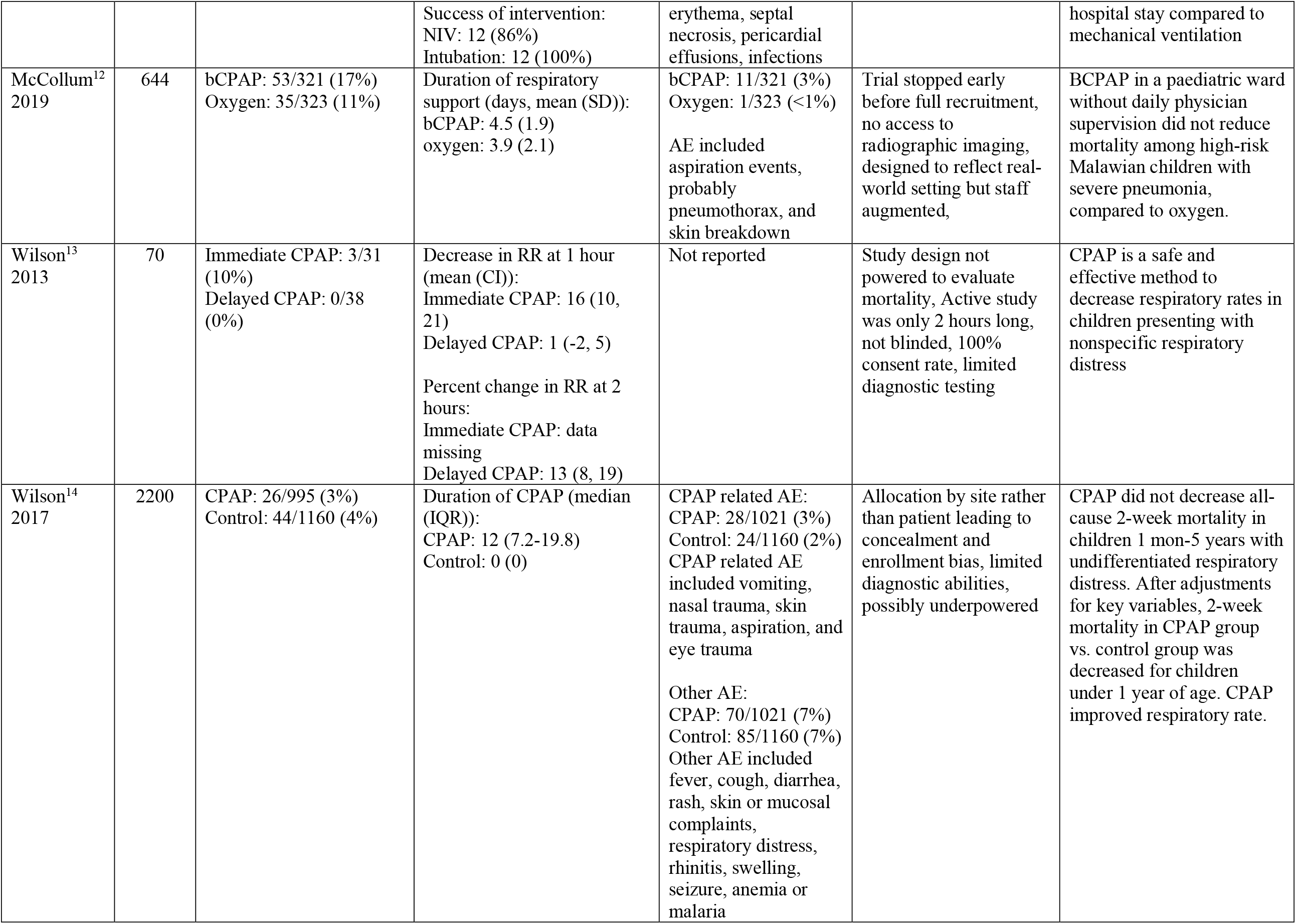
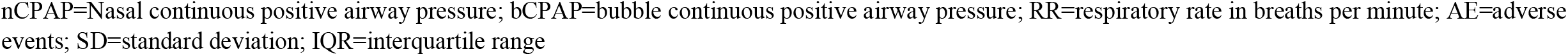
Outcomes for Randomized Control Trials

**Table 3b:** Outcomes for non-randomized control trials

| Author, year | Total Sample Size | Mortality | Additional Findings | Adverse Events | Reported Limitations | Reported Conclusions |
| --- | --- | --- | --- | --- | --- | --- |
| Balfour-Lynn<sup>16</sup><br>2014 | 106 | 2 (2%) | N/A | 0 (0%) | Possibility of missing data | NIPPV can be a simple and cost-effective way to treat patients with acute respiratory failure |
| Bjorklund<sup>17</sup><br>2019 | 83 | 8 (10%) | Patients with severe illness based on Tal score:<br>0 hours: 64/83 (77%)<br>2 hours: 12/83 (15%) | Severe: 0<br>Mild: 5 (6%)<br>Mild AE included nasal tissue irritation and abdominal distension | Evaluations for complications based only on clinical exam, not powered to evaluate effectiveness, differences in pre-trial and trial patients | SEAL-bCPAP is safe for treatment of respiratory distress in non-neonatal children in LMIC with a trend toward decreased mortality. |
| Bonora<sup>18</sup><br>2011 | 154 | Avoided intubation: 3.8%<br>Required intubation: 38.8% | No need for intubation: 80/154 (52%)<br>Duration of NIV (days, median (IQR)):<br>Avoided intubation: 4 (2.25-6)<br>Required intubation: 2 (1-4)<br>Duration of hospital stay (days, median (IQR)):<br>Avoided intubation: 6 (5-9)<br>Required intubation: 13 (9-24) | Skin breakdown noted but number of adverse events not reported | Retrospective study design with no control group, no rigid protocol to determine when therapies should be escalated or discontinued | NIV avoided mechanical ventilation in a high proportion of children. |
| Brown<sup>27</sup><br>2013 | 1 | 0 (0%) | Duration of bCPAP: 4 days<br>Duration of hospital stay: 6 days | 0 (0%) | Limitations not reported | A low-cost bCPAP could reduce child mortality in Africa. |
| Figueroa<sup>19</sup><br>2017 | 120 | Not reported | Success of bCPAP: 72%<br>Duration of bCPAP (hrs, mean (CI)): 75 (65-85) | 4 (3%)<br>Complications included abdominal | Limitations not reported | A reduction in respiratory rate, heart rate and TAL scores at 2 hours after starting |
|  |  |  | Duration of ICU stay (days, mean (CI)): 10 (9-11) | bloating and pneumothorax |  | intervention were predictors of success |
| Ghiggi<sup>20</sup><br>2000 | 42 | 2 (5%) | Duration of nasopharyngeal CPAP (days, mean (SD)): 4.12 (3.71)<br>Need for mechanical ventilation: 13/42 (31%) | 8 (19%)<br>Complications included tube obstructions and apnea due to excessive sedation | Small sample size | Nasopharyngeal CPAP was useful to avoid mechanical ventilation |
| Kinikar<sup>21</sup><br>2011 | 36 | 0 (0%) | Duration of ICU stay (days, median (range)): 2 (2-5)<br>Duration of hospital stay (days, median (range)): 7 (6-11)<br>Decrease in mean RR after 6 hours:<br>H1N1 positive: 20<br>H1N1 negative: 17 | 0 (0%) | Limitations not reported | Indigenous NB-CPAP improves hypoxemia and signs and symptoms in hemodynamically stable children with acute respiratory failure due to influenza-like injury |
| Lum<sup>22</sup><br>2011 | 129 | 19 (15%) | Duration of NIV (days, median (IQR)): 4 (2-8)<br>Duration of PICU stay (days, median (IQR)): 4.5 (2-9)<br>Avoided mechanical ventilation for $\geq 5$ days: 98 (76%) | 29 (22%)<br>Complications included pneumonia while on NIV, pressure from mask, and problems with mask fitting | Not an RCT, no routine use of blood gas sampling, shortage of NIV machines | NIV represents a viable strategy that provided effective respiratory support and prevented intubation in majority of patients. |
| Machen<sup>23</sup><br>2015 | 79 | 23 (29%) | Duration of CPAP (days, mean): 3.12<br>Duration of hospitalization (days, mean): 8.41<br>Had lower RISC score after 24 hours: 63 (80%) | Not reported | Clinical diagnoses could have led to misclassification | BCPAP was most beneficial to patients with bronchiolitis. |
| McCollum<sup>28</sup><br>2011 | 1 | 0 (0%) | Duration of bCPAP (days): 7 | 0 (0%) | Limitations not reported | BCPAP was successful in treating an infant with PJP pneumonia secondary to HIV infection |
| Myers<sup>24</sup><br>2019 | 117 | 38 (33%) | Required intubation:<br>15/115 (13%)<br>Duration of treatment<br>(hours, median (IQR)): 24<br>(24-60) | 13 (11%)<br>Complications<br>included blocked<br>nostrils or nasal<br>prongs, interruption<br>of oxygen supply,<br>nasal septum lesions,<br>and aspiration | Observational study<br>design, small sample<br>size, limited human<br>resources and some<br>missing data points, | It is feasible to use<br>bCPAP in the hospital<br>management of<br>critically ill children in<br>resource-limited<br>settings. |
| Pulsan<sup>25</sup><br>2019 | 64 | 35 (55%) | RDS (mean (IQR)):<br>Pre-CPAP: 11 (10-12)<br>1 hour: 9 (8-11)<br>84 hours: 6.5 (6-8) | Not reported | Observational study<br>design, bCPAP only<br>used when oxygen failed | BCPAP improves<br>oxygenation and<br>reduces respiratory<br>distress in some<br>children but children<br>with comorbidities<br>continue to do poorly |
| Walk<sup>26</sup><br>2016 | 77 | 36 (47%) | Duration of treatment<br>(days, median (IQR)): 3 (3-5) | 13 (17%) | Non-randomized and<br>uncontrolled, small<br>sample size,<br>understaffing, missing<br>vital sign data | BCPAP can be feasibly<br>implemented into a<br>tertiary African<br>hospital with high-risk<br>patients |

nCPAP=Nasal continuous positive airway pressure; bCPAP=bubble continuous positive airway pressure; RR=respiratory rate in breaths per minute; AE=adverse events; SD=standard deviation; IQR=interquartile range

**Figure 1:**
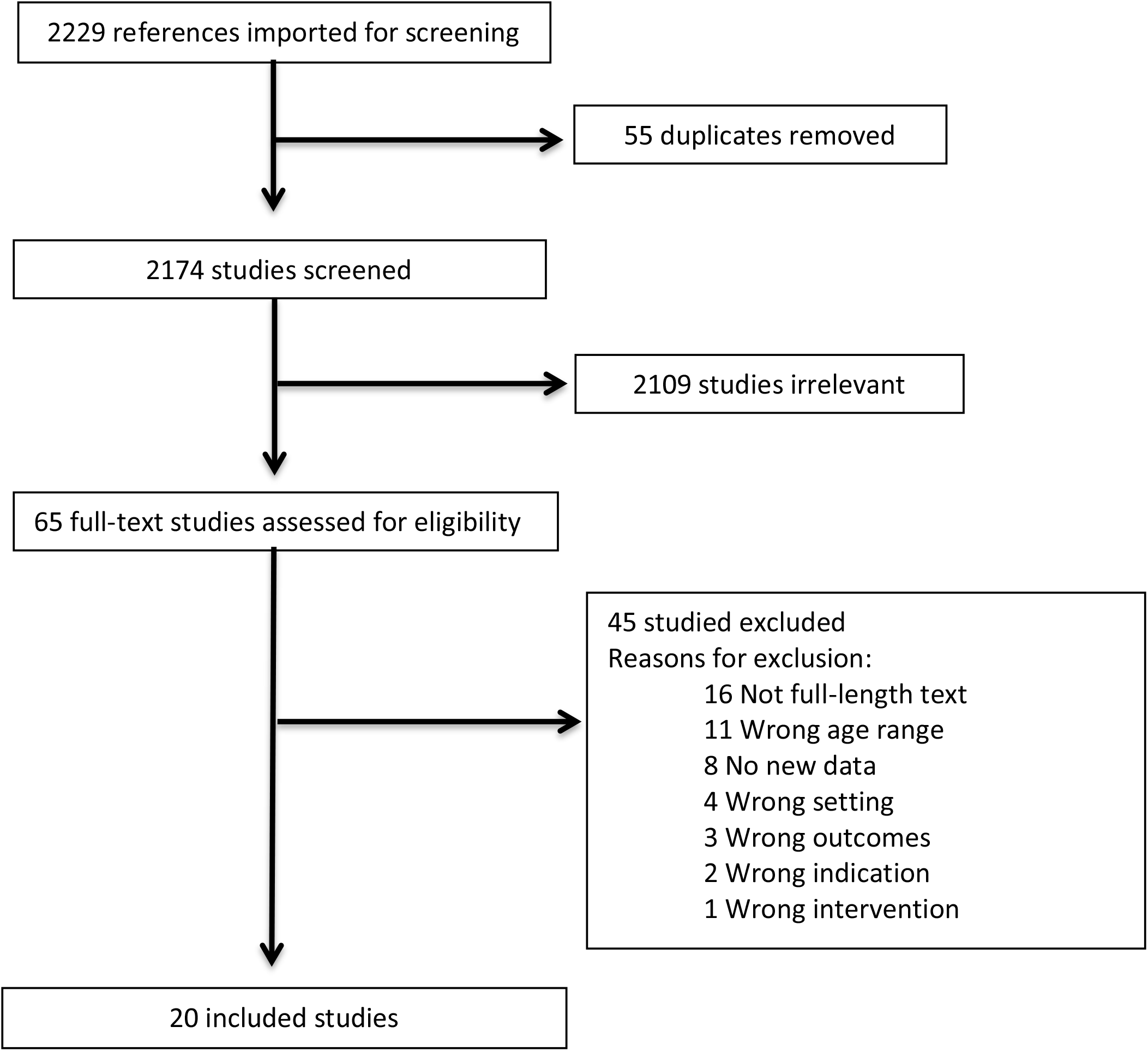
Study Selection.

Sixteen studies were carried out at tertiary referral or provincial hospitals. Of these, thirteen were specifically in intensive care units or other dedicated high acuity units or “zones”. Four studies, including the Malawi ^12^ and Ghana trials^14^ were carried out at district hospitals on the general pediatric ward. The Ghana trial had twice daily physician rounds while the Malawi trial was conducted by non-physician clinical staff with every-other-week pediatrician oversight.

Mortality was the primary endpoint for seven of the eighteen studies that reported it. For the five RCTs, mortality rates varied substantially, ranging from 0 – 22%. Mortality rate or treatment failure served as primary endpoints for the RCTs. One of the first RCTs, the Bangladesh trial, reported ‘treatment failure’ as the primary endpoint. CPAP was delivered in an ICU setting under supervision of a pediatric intensive care physician. Children who received bCPAP compared to low-flow oxygen had a significantly lower risk of treatment failure (RR 0.27, 99.7% CI 0.07 – 0.99; p=0.0026) and mortality (4% bCPAP versus 15% low-flow oxygen: RR 0.25, 95% CI 0.07-0.89; p=0.022).^10^ The study was stopped early by the data safety monitoring board (DSMB) for benefit. A second RCT, the Ghana trial, used a crossover design in which CPAP was available at one hospital at a time, while the other hospital served as the control.^14^ Children at the intervention hospital received CPAP and at both hospitals, supplemental oxygen was provided as needed to maintain appropriate oxygenation. The proportion of children in the control arm that received oxygen was not reported. This trial found no decrease in all-cause mortality between the CPAP and control arms (3% and 4% respectively; RR 0.67, 95% CI 0.42 – 1.08; p=0.11). However, an analysis adjusted for site, time and clinical variables, demonstrated decreased mortality for children under 1 year of age (3% CPAP and 7% control; RR 0.40, 95% CI 0.19 – 0.82; p=0.01).^14^ One RCT, the Malawi trial, compared bCPAP to low flow oxygen and found higher mortality in the bCPAP arm (17% and 11% respectively, RR 1.52; 95% CI 1.02 – 2.27; p=0.036).^12^ This study was stopped early due to both futility and the possibility of harm from bCPAP. In a small open, prospective RCT from Vietnam involving 37 children with respiratory distress due to dengue sock syndrome, 18 received nCPAP of 6 cmH2O and 19 received oxygen by facemask (6 – 8 LPM). Respiratory rate significantly improved in the nCPAP group after 30 minutes of treatment compared to control (p<0.05) and there was less treatment failure (22% nCPAP vs 68% control, p<0.01).^9^

Among the eleven observational LMIC studies, mortality rates for patients treated with NIV ranged from 0-55%. Four of these studies, all of which took place at tertiary hospitals, reported mortality rates >30%.^18,24-26^ Mortality was the primary endpoint for five prospective observational studies and was 2%^16^, 10%^17^, 29%^23^, 33%^24^, and 47%.^26^ Multiple comorbidities likely play a detrimental role in outcomes, as evidenced by the study with the highest mortality rate (47%), which showed much lower mortality rates among lower risk patients (18% rate in HIV-uninfected and only single organ failure).^26^ Similarly, the study with a reported 33% all-cause mortality among CPAP recipients reported no deaths among HIV-uninfected patients with very severe pneumonia and single organ failure.^24^ In this study, mortality was highest among patients with multiorgan failure (55%), severe malnutrition (64%), and HIV infection or exposure (55%). Younger patients fared better than older: patients aged 0 – 2 months had 93% lower odds of death or intubation than patients ≥60 months of age (OR 0.07, 95% CI 0.00-1.02; p=0.05).^26^ Despite a wide range in mortality, all five studies concluded that NIV can be beneficial in the treatment of respiratory distress in some children.

Fifteen studies reported on non-fatal adverse events (AEs). Six of these reported no AEs. Rates of AEs in the other seven studies ranged from 3-22%. One study reported a total AE rate of 79% including infections.^15^ When infections were excluded, the AE rate decreased to 22%. Most AEs were mild and included trauma to the nasal septum, skin, and eyes, vomiting, and abdominal distension.^12,14,15,17-19,24^ However, a few serious AEs including aspiration and pneumothorax were reported.^12,14,19^ Three studies reported AEs related to device malfunction, including poor mask fit, CPAP tube obstruction and disruption of the oxygen supply.^20,22,24^

### Risk of Bias Assessment

Seven studies were evaluated using the Cochrane GRADE criteria (Figure 2a). Due to the inability to blind which respiratory therapy a child is receiving, none of the studies were blinded from participants, personnel, or outcome assessors. One study was not randomized^15^ and another RCT utilized a cluster design and randomized at the hospital, rather than patient, level.^13^ All seven studies had low risk of incomplete data or reporting bias.

**Figure 2a:**
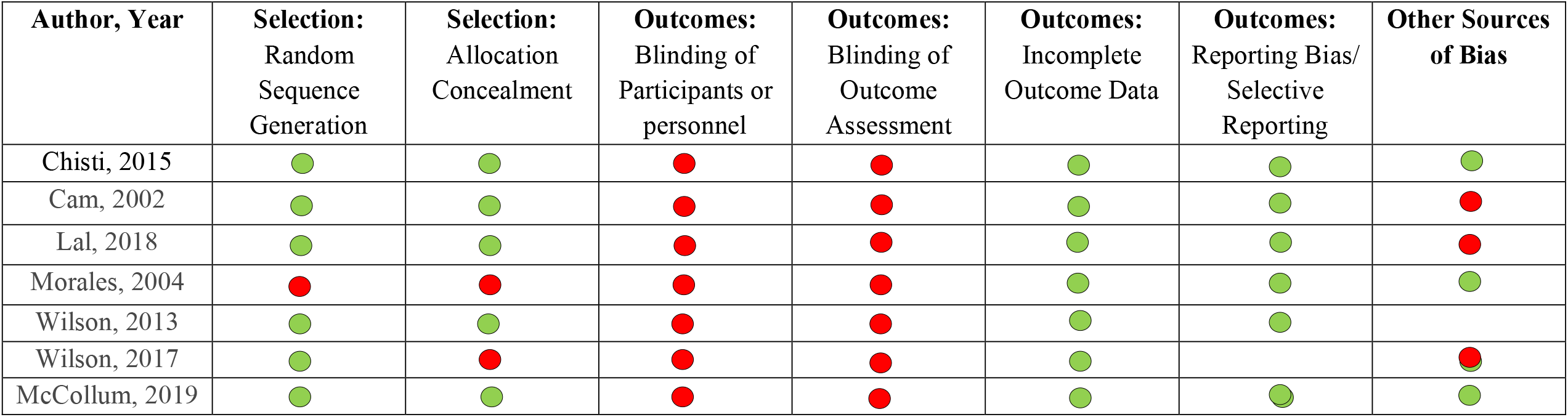
Risk of Bias Assessment for RCT and Prospective Comparative Studies using GRADE

Thirteen studies were evaluated using the criteria proposed by Murad et al.(Figure 2b).^8^ Five studies had unclear or high risk of selection bias due to inconclusive reporting of which patients were included and if all patients were captured.^16,23,25,27,28^ All studies were considered low risk of ascertainment bias, which assesses if exposures and outcomes were adequately ascertained. Due to observational study design, 10/13 studies were considered unclear or high risk of causality bias. Risk of causality bias was assigned based on potential alternate causes, presence of a challenge/rechallenge phenomenon, and appropriate duration of follow up.^8^

**Figure 2b:**
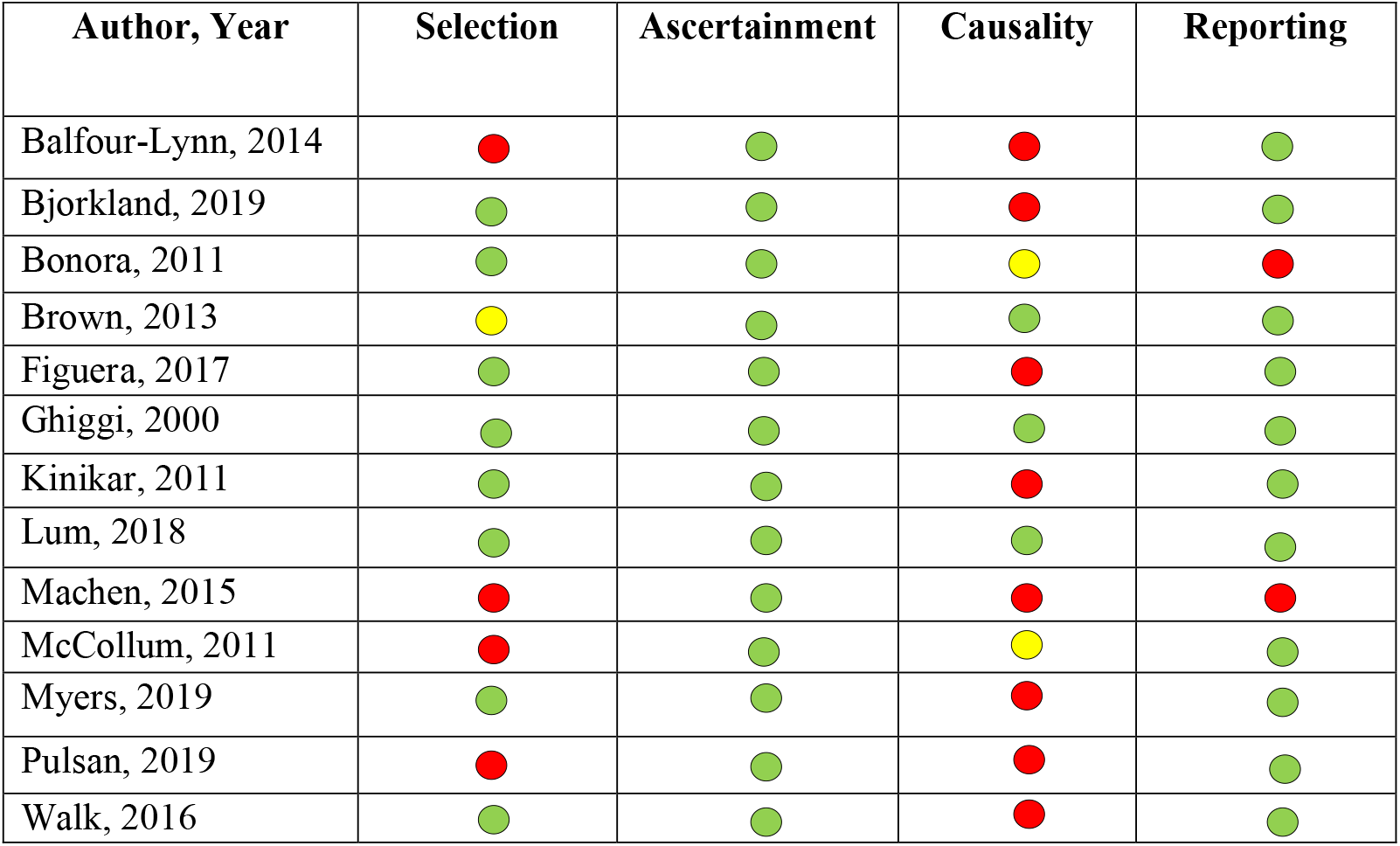
Risk of Bias Assessment for non-comparison studies including case-control, observational studies, and case studies

## Discussion

We completed a systematic review of all published articles related to the efficacy, effectiveness, and safety of CPAP in children between 1 month and 15 years of age in LMICs. Overall, we found that the evidence of efficacy is mixed regarding the use of NIV in children with respiratory failure in LMIC. In total, three adequately sized RCTs demonstrated inconsistent findings on the efficacy of CPAP compared to other respiratory modalities. The additional published reports include varying levels of benefit and mortality, making an assessment regarding effectiveness difficult. Our review of the literature suggests that CPAP for non-neonatal pediatric patients should not be administered outside well controlled, monitored environments under direct physician supervision until further evidence is available.

The three RCTs that included larger sample sizes are worth placing into context with each other given none provides conclusive evidence of efficacy for the use or avoidance of CPAP. While the Bangladesh trial was stopped early for benefits, some argue that the trial’s closure did not adhere to pre-defined stopping rules.^29^ Since the trial’s setting was in an ICU, extrapolating care to other more typical resource constrained LMIC environments remains difficult. This however does not deter from the findings of the study that CPAP may be safely used in an ICU setting with direct physician oversight. The Ghana trial did not demonstrate any difference in its primary outcome of mortality when considering all participants. However, for children less than 1 year of age, the Ghana trial did demonstrate a mortality benefit for CPAP as compared to the control group who received low-flow oxygen during exploratory analyses. It is also unclear what proportion of control patients received low flow oxygen, but the low baseline hypoxemia prevalence suggests it could have been a minority of children. This is likely an important methodological difference from the other two RCTs that administered oxygen to all control patients. Of note, the study was also conducted under physician oversight in a district hospital’s emergency department dedicated beds. Finally, the Malawi trial was stopped early for both futility and potential harm from CPAP as compared to low-flow nasal cannula. The Malawi trial targeted and enrolled distinctly sicker children as all patients had to have at least one comorbidity or hypoxemia. Further, the trial was conducted in a district pediatric ward hospital without daily physician oversight.

When reviewing all AEs, excluding mortality, we found that AEs were relatively rare, and most were minor such as nasal trauma. Significant AEs were reported rarely and included aspiration and pneumothorax. Investigators from the Malawi trial postulate that aspiration may have been a factor in its results. Pneumothorax, while a theoretical adverse event, appears rare when using CPAP. Taken together, these results do offer guidance on best practices for CPAP use in LMICs for children older than 1 month.

Given mixed evidence, further research with well-designed RCTs will be critical to advance our understanding of efficacy and feasibility. As more pediatric services in LMICs begin to implement CPAP in their respective units, equipoise will begin to wane, making it difficult to build a strong evidence base built on RCTs. Furthermore, well-developed management approaches need to be developed given the potential physiologic complexity of CPAP treatment, taking into consideration resource constraints and related ethical dilemmas. For example, if oxygen concentrators are used in a bCPAP system to drive gas flow, then one child occupies one full concentrator, when flow from the same concentrator could potentially be used to simultaneously support upwards of 5 children requiring low-flow oxygen. A strong understanding of which patient populations will derive maximum benefit from CPAP is critical before diverting possibly life-saving resources away from other children.

Importantly, we recognize that despite mixed evidence, CPAP is currently being implemented as a respiratory modality in LMICs. While we argue for further research prior to wider implementation of CPAP, we offer some guidance. Based on the evidence thus far available, we first suggest that CPAP be used only with direct physician oversight. This is borne out by the three major RCTs discussed above, reflecting that CPAP and its surrounding care is complex. At this time evidence suggests that relying on nurse driven protocols is insufficient to safely use CPAP for non-neonatal pediatric care. It is suggested that medical facilities aiming to provide CPAP maintain a high-level of training to cultivate a robust depth of knowledge of CPAP use and principles; providers should know how and when to initiate, escalate or deescalate, and stop CPAP support, using well-developed and widely-accepted tools.

Second, CPAP should be limited at this time to children less than 1 year of age. Currently, there are no compelling data to support its use in older children. Third, CPAP is best used in an intensive care, high dependency, or dedicated unit with greater staff to patient ratios. Fourth, aspiration remains a possible risk and feeding, if at all, should be done with caution while patients are on CPAP. Finally, given these mixed efficacy and effectiveness data we strongly recommend that in settings moving forward with CPAP implementation a parallel monitoring and evaluation system to track and annually report on cohorts of CPAP patients is required. Key indicators to include are the following: age, sex, underlying medical conditions, primary admission diagnosis, final diagnosis, final secondary diagnoses, admission SpO2, admission vital signs (temperature, heart rate, blood pressure, respiratory rate, mental status), CPAP nasal interface (nasal or full face mask, prongs, other), CPAP system, CPAP flow source, oxygen source, feeding status, nasogastric tube use, intravenous fluid use, duration of CPAP treatment, duration of hospitalization, hospital outcome.

Our systematic review demonstrates that current data for CPAP is mixed and does not offer conclusive evidence for CPAP in LMICs. If CPAP is to be used as a respiratory modality in LMICs it should be used with caution, under strict physician supervision in a critical care setting with high provider-to-patient ratios, and with standardized annual reporting of key indicators for full programmatic transparency in order to optimize patient safety.

## Data Availability

The corresponding author has full access to the data which is available upon request.

## Contributors

KS had full access to all the data in the study and takes full responsibility for the integrity of the data and the accuracy of the data analysis. KS and EM conceived the study idea and designed the study; KS, and EM with the help of multiple medical librarians created the search terms and conducted the database searches; KS and PH extracted and analyzed the data; KS, AS, PH, RC, and EM drafted and revised the manuscript. All authors made substantial contributions to the interpretation of results, critical revision of the manuscript, and approved the final version of the manuscript.

## Acknowledgments

We would like to thank Ann Farrell, Peggy Murphy, and the medical library staff at Mayo Clinic and Lurie Children’s Hospital who helped with search criteria and preformed the searches.

## Panel: Research in context

### Evidence before this study

- Non-invasive ventilation (NIV) is safe and effective for the treatment of respiratory distress in pediatric patients in high income countries
- NIV is safe and effective for the management of newborns with respiratory distress in low and middle income (LMIC) countries

### Added value of this study

- Analysis of all available research evaluating the safety and efficacy of NIV in LMICs for non-neonatal pediatric patients

### Implications of all the available evidence

- Current data for CPAP is mixed and does not offer conclusive evidence for CPAP in LMICs
- If CPAP is to be used as a respiratory modality in LMICs it should be used with caution and under strict care

